# The Trend of Ischemic Evaluation, Intervention, and In-Hospital Mortality and Outcomes among Patients with Acute Myocardial Infarction and Atrial Fibrillation

**DOI:** 10.1101/2023.12.15.23300053

**Authors:** Mirza Faris Ali Baig

## Abstract

**Background:** Atrial fibrillation (AF) is associated with increased cardiovascular mortality. Data regarding the relationship between coronary artery disease (CAD) and AF is mixed. It is uncertain if AF directly increases the risk for future coronary events and if such patients are appropriately evaluated for CAD.

**Methods:** This is a cross-sectional study performed on hospitalized patients with AMI and concurrent AF in 2019 using National Inpatient Sample from HCUP. Patients with missing information and type II non-ST elevation myocardial infarction (NSTEMI) were excluded. Using STATA 18, In-hospital mortality, ischemic evaluation, percutaneous treatment, rates of ventricular tachycardiac (VT), ventricular fibrillation (VF), cardiogenic shock, cardiac arrest, average length of stay (LOS), and total hospitalization charges were studied. Regression models were used for data analyses.

**Results:** A total of 600,645 patients met inclusion criteria (219,660 females [36.5%], 428,755 Caucasian [71%], 65,870 African American [10.9%], 51,155 Hispanic [8.5%]; mean [SD] age, 66.7 [0.5] years), including 166,680 (28%) STEMI and 433,965 (72%) NSTEMI patients. 109,520 (18%) patients with AMI had AF. For patients with AMI and AF, the adjusted odds of mortality increased by 23% (adjusted Odds ratio [aOR], 1.23; CI, 1.15-1.32; p<0.001). AF patients were less likely to undergo ischemic evaluation (aOR, 0.77; CI, 0.74-0.80; p<0.001) and ischemic intervention (aOR, 0.64; CI, 0.62-0.66; p<0.001). AF patients had higher odds of VT (aOR, 1.41; CI, 1.33-1.49; p<0.001), VF (aOR, 1.44; CI, 1.33-1.57; p<0.001), cardiogenic shock (aOR, 1.43; CI, 1.35-1.52; p<0.001), and cardiac arrest (aOR, 1.35; CI, 1.24-1.47; p<0.001). AF patients had longer LOS (mean, 1.39; SCD, 1.29-1.48; p<0.001) and higher total hospital charges (mean $22,188; 19,311-25,064, p<0.001).

**Conclusion:** AF was independently associated with increased mortality in patients admitted with AMI. AF was associated with higher rates of cardiac complications. Patients with AF were less likely to receive ischemic evaluation or percutaneous intervention and had overall higher healthcare resource utilization. This study encourages AF to be viewed as an independent risk factor for CAD and suggests more efforts to diagnose CAD in such patients.

**Clinical Perspective:** *What is new:*

- Patients with acute myocardial infarction and atrial fibrillation have higher odds of mortality.
- AMI patients with AF are subjected to lesser odds of undergoing ischemic evaluation and intervention.
- Healthcare resource utilization is higher in this cohort.

*What are the clinical implications:*

- AF should be considered as an independent risk factor for increased mortality in AMI.
- Early ischemic evaluation should be considered to diagnose coronary artery disease in this cohort.
- Increased awareness to recognize all risk factors of coronary artery disease.

## Introduction

Atrial fibrillation (AF) is the most common cardiac arrhythmia globally.^1^ The prevalence of AF in the US is at least 3-6 million and is projected to increase to 6-16 million by 2050.^2^ The lifetime risk of acquiring AF is estimated to be 33%.^3^ AF is known to increase mortality from sudden cardiac death, congestive heart failure, and stroke;^4^ therefore, it is not surprising that healthcare costs associated with AF are significantly higher.^5^ Coronary artery disease (CAD), on the other hand, is the most common cardiovascular disease.^6^ It is the leading cause of mortality in the US, with approximately 610,000 deaths annually,^7^ and the third leading cause of death worldwide, with 17.8 million deaths annually.^8, 9^ Healthcare costs related to CAD are estimated to be more than US$200 billion annually.^10^ Due to the colossal healthcare burden, considerable efforts have been made to understand the underlying risk factors. These risk factors are segregated into two large categories: modifiable and non-modifiable risk factors. The non-modifiable risk factors include age, gender, family history, and certain ethnicities. The known modifiable risk factors include hypertension, diabetes mellitus, hyperlipidemia, obesity, smoking, sedentary lifestyle, and stress.^11^ Significant efforts are made to discover novel risk factors, aiming to formulate additional guidelines to alleviate substantial healthcare costs and mortality rates.

While both diseases are known to share a common risk factor profile, the intricate relationship between the two is still largely unclear. It is reported that CAD is a risk factor for AF;^12^ however, studies have shown mixed results about AF being an independent risk modifier for CAD, with some proposing direct causality,^13^ whereas others suggesting no causal relationship between AF and risk of CAD.^14^ It is not uncommon in practice for patients to have both AF and CAD. AF is known to have a negative prognostic impact on CAD patients;^15^ yet, it remains unclear if the increased mortality is due to increased coronary ischemic events or from unfavorable hemodynamic effects, such as loss of atrial kick, high ventricular rate, and decreased cardiac output.^16^

Several studies have attempted to understand the impact of both diseases. While previous studies have explored the associated poor outcomes, no study has directly examined the pattern of diagnostic workup, intervention, and cardiac outcomes in this sub-population. This study aims to investigate the effect of AF on acute myocardial infarction (AMI) in terms of in-hospital mortality, serious cardiac complications, and hospital resource utilization and explore the national trend of ischemic evaluation and treatment in patients with both AMI and AF.

## Methods

This is a retrospective cohort study of adult patients hospitalized in the United States with the diagnosis of AMI. The study used the patient cohort from the National Inpatient Sample (NIS) database from January 1, 2019 to December 31, 2019. The NIS is a part of the Healthcare Cost and Utilization Project (HCUP), maintained by the Agency for Healthcare Research and Quality (AHRQ).^17^ NIS is sampled from the State Inpatient Databases (SID), containing information on all hospital stays regardless of the payer source. NIS 2019 sampling frame included data from 49 statewide organizations, estimated to have 97% of discharges from non-federal short-term US hospitals, covering 98% of the US population. NIS collects 20% of the stratified sample of discharge records from all HCUP-participating US community hospitals, excluding long-term acute care facilities and rehabilitation centers. Discharge weights are provided to produce national estimates, approximating 35 million discharges when weights are applied. This allows NIS to provide reliable information about cardiovascular diseases.

### Study patients

Patients who were adults (≥18) and admitted non-electively were included. The ICD-10 CM codes were used to identify patients hospitalized for ST-elevation myocardial infarction (STEMI) and non-ST elevation myocardial infarction (NSTEMI). These patients were then accumulated under the diagnosis of AMI for data analysis. The ICD-10 codes used are listed in the attached Appendix. Patients with the diagnosis of NSTEMI type II or acute myocardial injury due to secondary causes or a procedural complication were excluded. Patients with missing information were also excluded from the analysis.

### Study variables

ICD-10 procedure codes were used to identify patients who underwent diagnostic evaluation for coronary artery disease. Diagnostic cardiac angiograms and non-invasive cardiac stress tests were identified using the ICD-10 procedure codes and gathered under a single variable.

Percutaneous coronary intervention (PCI) procedure codes were used to identify patients who underwent any percutaneous intervention (see Appendix). Secondary diagnosis codes were used to identify AF patients. Separate variables were created for all other comorbid conditions. Length of stay (LOS) and total hospitalization charges are provided within the NIS data for each hospital admission. Covariates included in the study were age, gender, race, baseline comorbidity status using the Charlson Comorbidity Index, median annual income in the patient’s zip code, geographic region of the United States (Northeast, Midwest, West or South), hospital location (urban vs rural), hospital teaching status, hospital bed size, primary insurance, hypertension, diabetes mellitus, hyperlipidemia, smoking status, obesity, history of coronary artery disease, history of myocardial infarction, history of percutaneous coronary intervention, systolic heart failure, diastolic heart failure, combined systolic and diastolic heart failure, history of peripheral vascular disease, chronic pulmonary disease, pulmonary hypertension, obstructive sleep apnea, acute kidney injury, chronic kidney disease, chronic liver disease, sepsis, septic shock, alcohol use, cocaine, amphetamine use and anemia of chronic disease.

### Outcomes

The primary outcome was in-hospital mortality. Secondary outcomes were ischemic evaluation, percutaneous ischemic intervention, rates of ventricular tachycardia (VT), ventricular fibrillation (VF), cardiogenic shock, cardiac arrest, length of stay (LOS), and total hospitalization charges. NIS provides in-hospital mortality, LOS, and total hospital charges. Separate variables were created for other secondary outcomes.

### Statistical Analysis

STATA 18.0 was used to analyze the results. NIS is generated using a complex sampling design, including stratification, clustering, and weighting. STATA can run analyses to provide nationally representative unbiased results and generate p values. Univariable and multivariable linear regression analyses were used to calculate means for continuous variables. Univariable logistic regression analysis was used to calculate unadjusted odds ratios (ORs) for categorical and dichotomous variables. A separate univariable logistic regression analysis was performed to isolate statistically significant variables with a p-value of less than 0.02. Those variables were then included in the multivariate logistic regression analysis to adjust for potential confounders. Another analysis was conducted incorporating variables with p-values between 0.02 and 0.05, along with those above 0.05 but known to be confounders in previous studies, to validate our findings.

Fisher exact test was used to compare proportions. Student t-test was used for continuous variables. All p-values obtained were two-sided. 0.05 was used as the threshold for statistical significance.

## Results

### Patient characteristics

A total of 35 million weighted discharges were included in the NIS 2019, of which 600,645 met the inclusion criteria of our study. 36% of the patients were females, and the predominant race was Caucasian (73%). 166,680 (28%) patients had a primary diagnosis of ST-elevation myocardial infarction (STEMI) and 433,965 (72%) with non-ST elevation myocardial infarction (NSTEMI). 109,520 (18%) patients had a secondary diagnosis of AF. Patients with the secondary diagnosis of AF were more likely to be older, Caucasian, and insured by Medicare compared to the patients without AF. Table 1 summarizes the baseline characteristics. The distribution of the female gender, Charlson comorbidity score, and other hospital-level parameters such as the US geographic region, hospital location, and hospital bed size were similar in both groups. Although these differences were statistically significant, the absolute differences were small.

**Table 1.** Patient characteristics.

| Patient characteristics | AMI with AF | AMI without AF | p-value |
| --- | --- | --- | --- |
| % of patients |  |  |  |
| Women, % | 37% | 36% | 0.0153 |
| Age, mean | 74 years (0.5) | 65 years (0.2) |  |
| Race/ethnicity, % |  |  | 0.000 |
| Caucasian | 80% | 72% |  |
| African American | 7.5% | 12% |  |
| Hispanic | 6.5% | 9.2% |  |
| Asian or Pacific Islander | 2.6% | 3% |  |
| Native American | 0.4% | 0.6% |  |
| Other | 2.3% | 3.1% |  |
| Charlson comorbidity index score, no. (%) |  |  |  |
| 0 | 0 | 0 | 0.000 |
| 1 | 11.6% | 26% |  |
| 2 | 19.5% | 26% |  |
| >3 | 69 | 48% |  |
| Median annual income in patients' zip code, % |  |  | 0.000 |
| \$1 - 45,999 | 28.6% | 31% | |
| \$46,000 - 58,999 | 27.2% | 26.3% | |
| \$59,000 - 78,999 | 25% | 24% | |
| > \$79,000 | 19.3% | 18.3% | |
| Insurance type, % |  |  | 0.000 |
| Medicare | 77% | 54% |  |
| Medicaid | 5% | 11% |  |
| Private HMO | 15% | 30% |  |
| Self-pay | 2.3% | 5.7% |  |
| Hospital characteristics |  |  |  |
| Hospital region, % |  |  | 0.000 |
| Northeast | 17% | 17% |  |
| Midwest | 24% | 22% |  |
| South | 39% | 41% |  |
| West | 20% | 19% |  |
| Hospital bed size, % |  |  |  |
| Small | 19% | 19% |  |
| Medium | 30% | 30% | 0.3233 |
| Large | 51% | 50% |  |
| Urban location | 93% | 93% | 0.853 |
| Hospital teaching status | 74 | 74 | 0.0903 |
| AMI, Acute myocardial infarction; AF, atrial fibrillation. Actual numbers not provided as the sample size was above 10,000 |  |  |  |

### In-hospital mortality

The overall in-hospital mortality rate for AMI was 4.5%. The difference in mortality between patients with and without AF is presented in Table 2. AF independently predicted increased odds of mortality (adjusted OR, 1.23; CI, 1.15-1.32; p<0.001) on univariable and multivariable analyses that adjusted for patient- and hospital-level confounders. In-hospital mortality rates for STEMI and NSTEMI were 7.8% and 3.2%, respectively. Similar associations were found for STEMI (adjusted OR, 1.23; CI, 1.09-1.38; p<0.001) and NSTEMI (adjusted OR, 1.32; CI, 1.21-1.45; p<0.001), as summarized in Table 3.

**Table 2.**
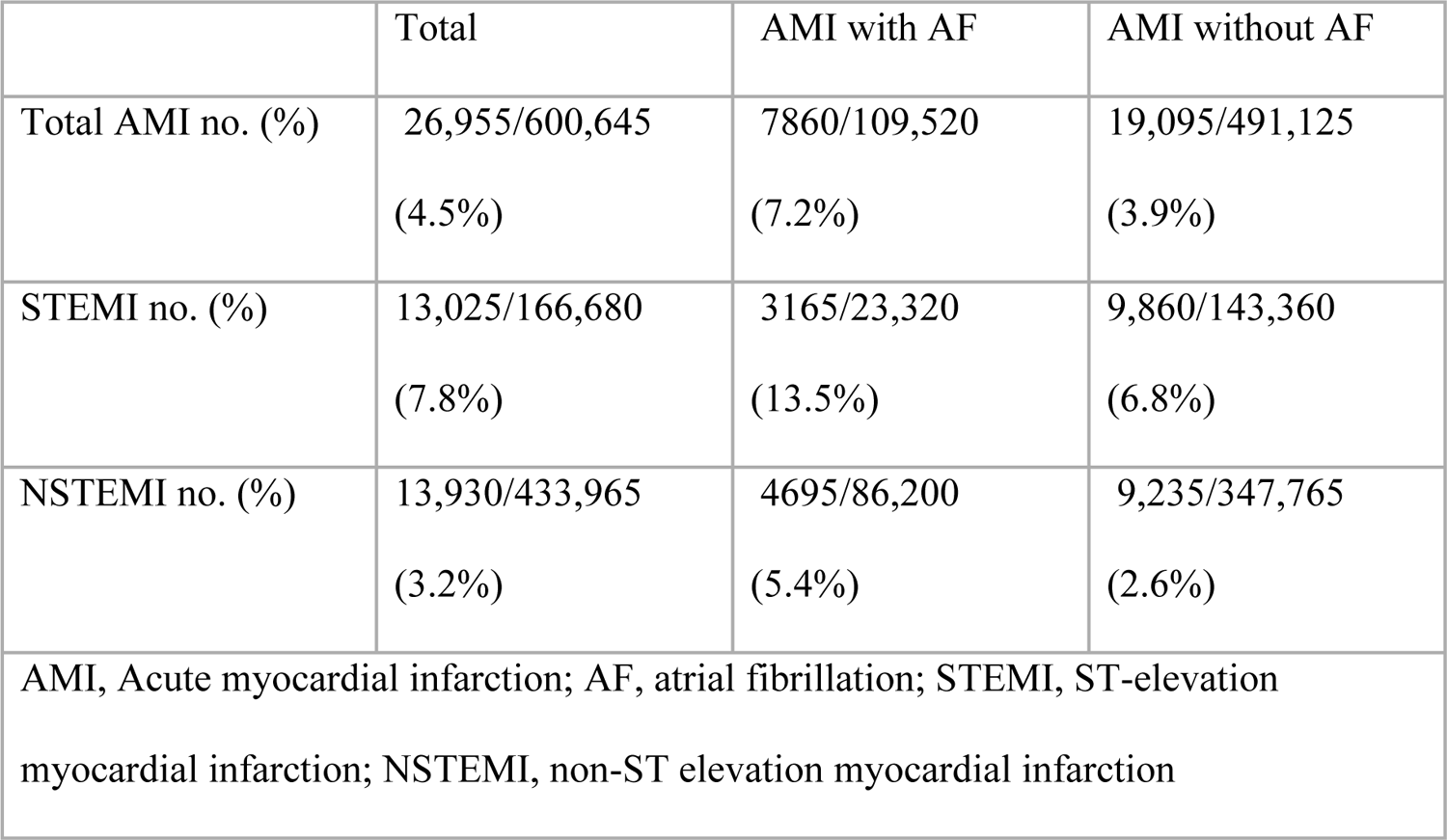
The difference in mortality between patients with and without AF.

**Table 3.** Adjusted and unadjusted odds of mortality in patients with AMI, STEMI, and NSTEMI and concomitant AF.

| Mortality | AMI |  | STEMI |  | NSTEMI |  |
| --- | --- | --- | --- | --- | --- | --- |
|  | Unadjusted | Adjusted | Unadjusted | Adjusted | Unadjusted | Adjusted |
|  | OR (95% CI) | OR (95% CI) | OR (95% CI) | OR (95% CI) | OR (95% CI) | OR (95% CI) |
|  | 1.91 (1.79-2.03) | 1.23 (1.15-1.32) | 2.12 (1.93-2.34) | 1.23 (1.09-1.38) | 2.11 (1.95-2.28) | 1.32 (1.21-1.45) |
| AMI, Acute myocardial infarction; AF, atrial fibrillation; STEMI, ST-elevation myocardial infarction; NSTEMI, non-ST elevation myocardial infarction |  |  |  |  |  |  |

### In-hospital ischemic evaluation

For patients admitted with AMI, the rate of in-hospital ischemic evaluation was 76%. Regarding in-hospital ischemic evaluation, AF patients were less likely to undergo ischemic evaluation with a diagnostic coronary angiogram or cardiac stress test after adjusting for confounders (adjusted OR, 0.77; CI, 0.74-0.80; p<0.001).

Ischemic evaluation was performed in 90% and 71% of patients with STEMI and NSTEMI admissions. As seen with AMI, patients with AF were less likely to undergo an ischemic evaluation than those without AF for STEMI (adjusted OR, 0.78; CI, 0.70-0.86; p<0.001) and NSTEMI (adjusted OR, 0.79; CI, 0.75-0.82; p<0.001). Like the primary outcome of mortality, we obtained similar results after including all the variables with p >0.02 but less than 0.05.

### In-hospital ischemic intervention

Among the patients admitted with AMI, 51.4% underwent PCI. 37.1% of patients with AF and AMI had PCI compared to 54.6% without AF. After univariable screening and multivariable logistic regression to neutralize potential confounders, AF patients were less likely to undergo PCI than patients without AF (adjusted OR, 0.64; CI, 0.62-0.67; p<0.001), detailed in Table 4. In a separate analysis, PCI was performed on 81.2% of all STEMI patients overall, though rates were lower for patients with AF (70%) than those without AF (83.1%). After adjusting for confounders, AF was an independent predictor of decreased odds of in-patient PCI (Adjusted OR, 0.65; CI, 0.59-0.70; p<0.001). 40% of patients admitted with NSTEMI underwent PCI. Patients diagnosed with AF had lower rates of PCI (28.2%) compared to patients without AF (43%). AF was again an independent predictor for lesser odds of in-patient ischemic intervention (Adjusted OR, 0.62; CI, 0.59-0.64; p<0.001).

**Table 4.**
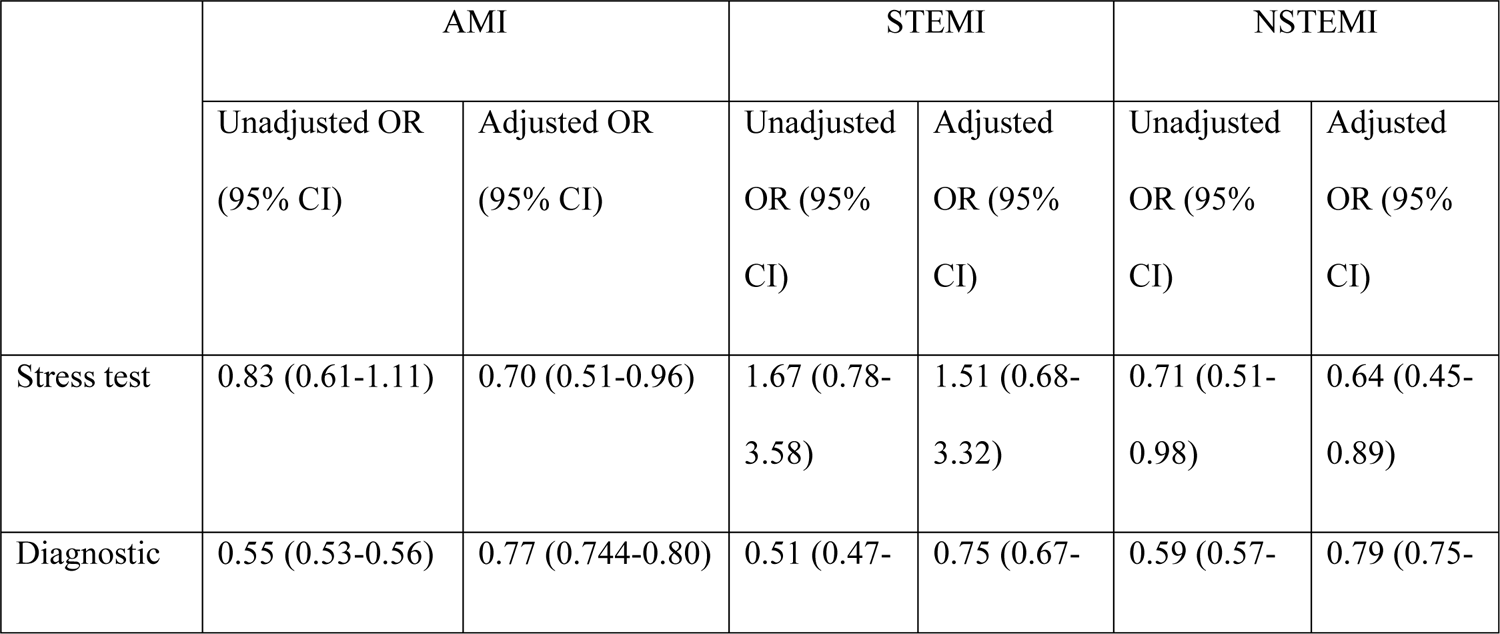
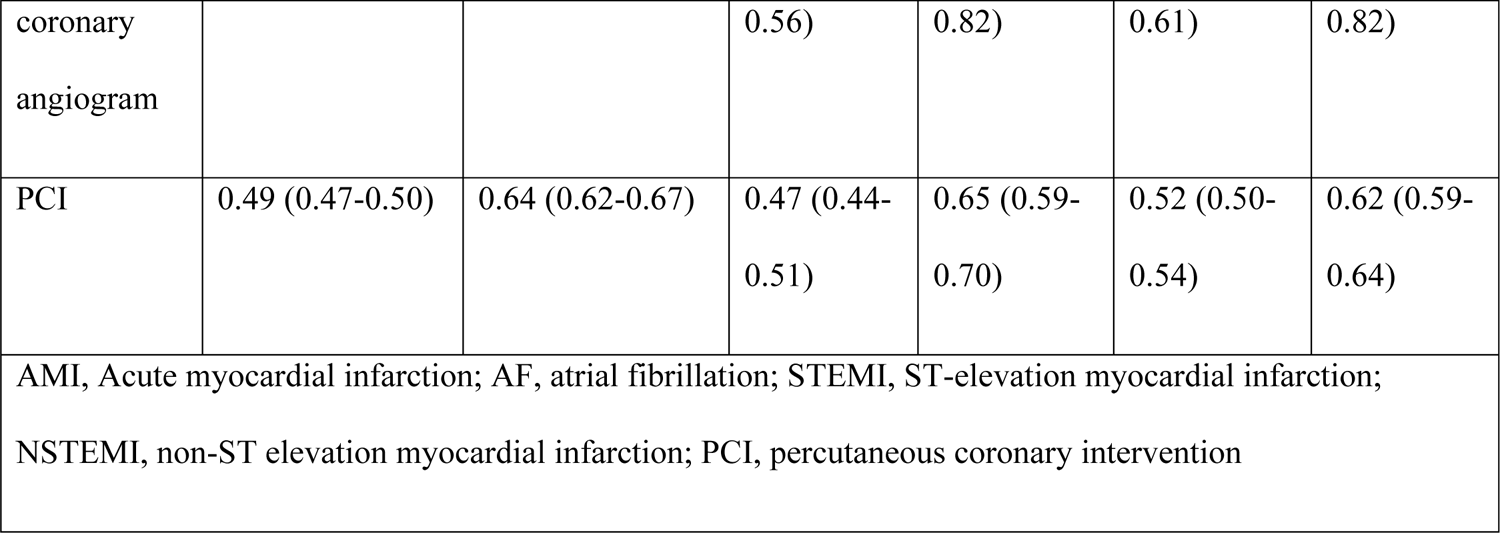
The unadjusted and adjusted Odds ratios of stress test, diagnostic coronary angiogram, and PCI in patients with AMI, STEMI, NSTEMI, and AF.

### Other important in-hospital cardiac outcomes

The study assessed the occurrence rates of in-hospital serious cardiac complications such as ventricular tachycardia (VT), ventricular fibrillation (VF), cardiogenic shock, and cardiac arrest. The rates of VT were 7.3%, 13.5%, and 4.9% for AMI, STEMI, and NSTEMI, respectively. Notably, these rates were elevated in individuals with AF, with 9.5% vs. 6.8% for AMI, 18.3% vs. 12.7% for STEMI, and 7.2% vs. 4.4% for NSTEMI.

Similarly, VF rates were 3.4% in AMI, 8.7% in STEMI, and 1.3% in NSTEMI. Again, AF was associated with higher rates (4.2% vs. 3.2% for AMI, 12.3% vs. 8.2% for STEMI, and 1.98% vs. 1.1% for NSTEMI). Incidences of cardiogenic shock were 6.7%, 14.1%, and 3.9% for AMI, STEMI, and NSTEMI, respectively. AF patients demonstrated elevated rates across all three groups (10.4% vs. 5.9% in AMI, 24.7% vs. 12.4% in STEMI, and 6.5% vs. 3.3% in NSTEMI). The rates of cardiac arrest were 2.9% in AMI, 5.9% in STEMI, and 1.7% in NSTEMI. Upon separate analysis based on AF status, higher cardiac arrest rates were observed in AF patients (4% vs. 2.7% in AMI, 9.1% vs. 5.4% in STEMI, and 2.6% vs. 1.5% in NSTEMI patients). As summarized in Table 5, AF independently predicted increased odds of VT, VF, cardiogenic shock, and cardiac arrest.

**Table 5.** Unadjusted and Adjusted Odds ratios for Ventricular tachycardia, Ventricular fibrillation, Cardiogenic shock, and Cardiac arrest in patients with AMI, STEMI, NSTEMI, and AF.

|  | AMI |  | STEMI |  | NSTEMI |  |
| --- | --- | --- | --- | --- | --- | --- |
|  | Unadjusted<br>OR (95% CI) | Adjusted<br>OR (95%<br>CI) | Unadjusted<br>OR (95%<br>CI) | Adjusted<br>OR (95%<br>CI) | Unadjusted<br>OR (95%<br>CI) | Adjusted OR<br>(95% CI) |
| Ventricular<br>tachycardia | 1.44 (1.36-1.52) | 1.41 (1.33-1.49) | 1.53 (1.41-1.66) | 1.44 (1.32-1.58) | 1.68 (1.57-1.80) | 1.44 (1.33-1.56) |
| Ventricular<br>fibrillation | 1.32 (1.22-1.42) | 1.44 (1.33-1.57) | 1.57 (1.43-1.73) | 1.50 (1.34-1.68) | 1.74 (1.52-1.98) | 1.51 (1.30-1.76) |
| Cardiogenic<br>shock | 1.83 (1.73-1.93) | 1.43 (1.35-1.52) | 2.31 (2.14-2.49) | 1.57 (1.43-1.73) | 2.05 (1.89-2.22) | 1.46 (1.33-1.59) |
| Cardiac arrest | 1.52 (1.41-1.64) | 1.35 (1.24-1.47) | 1.74 (1.56-1.94) | 1.50 (1.32-1.71) | 1.73 (1.54-1.94) | 1.43 (1.26-1.63) |
| AMI, Acute myocardial infarction; AF, atrial fibrillation; STEMI, ST-elevation myocardial infarction; NSTEMI, non-ST elevation myocardial infarction |  |  |  |  |  |  |

### Hospital length of stay and total hospitalization charges

Patients with AMI had a mean length of stay of 4.36 days, with associated hospitalization charges of $108,409; STEMI patients stayed 3.9 days, incurring charges of $126,896, while NSTEMI patients had a mean stay of 4.5 days with charges totaling $101,301. Following adjustments for patient- and hospital-level variables, individuals with AF exhibited significantly prolonged hospital stays and higher hospitalization charges compared to those without atrial fibrillation, as summarized in Table 6 below:

**Table 6.** Adjusted length of stay and total hospital charges for AMI, STEMI NSTEMI, and AF.

|  | AMI |  | STEMI |  | NSTEMI |  |
| --- | --- | --- | --- | --- | --- | --- |
|  | Unadjusted<br>OR (95%<br>CI) | Adjusted<br>OR (95%<br>CI) | Unadjusted<br>OR (95%<br>CI) | Adjusted<br>OR (95%<br>CI) | Unadjusted<br>OR (95%<br>CI) | Adjusted<br>OR (95%<br>CI) |
| LOS | 2.23 (2.12-2.33) | 1.39 (1.29-1.48) | 2.5 (2.28-2.72) | 1.49 (1.28-1.71) | 2.11 (2.00-2.22) | 1.34 (1.24-1.44) |
| Total<br>charges | 27,632<br>(24,445-30,818) | \$22,188<br>(19,311-25,064) | 39,063<br>(33,125-45,000) | \$22,577<br>(16,622-28,532) | 27,098<br>(23,817-30,379) | \$20,843<br>(17,898-23,788) |
| AMI, Acute myocardial infarction; AF, atrial fibrillation; STEMI, ST-elevation myocardial infarction; NSTEMI, non-ST elevation myocardial infarction; LOS, length of stay |  |  |  |  |  |  |

## Discussion

This study demonstrates that patients with AMI and concomitant AF had poorer outcomes than those without AF. This subpopulation was not only found to have increased mortality, but they were also less likely to undergo ischemic evaluation and treatment, with higher rates of cardiac complications, increased length of stay, and higher total hospitalization charges.

The interaction and causality relationship between CAD and AF is complex, and a large number of studies in the past have shown varied results. The prevalence of CAD in AF was previously estimated to be between 18-46.5%, whereas the prevalence of AF in CAD was low, from 0.5 to 5%.^12, 18^ In this study, the prevalence of CAD in AF was 40%, and the prevalence of AF in CAD was 29%. One plausible explanation for such divergence is that most previous studies captured longitudinal data and had fewer patients than this study. Previous research identified CAD as an independent risk factor for the development of AF,^19^ and early onset AF post-AMI is linked with a dismal prognosis.^20^ Studies have demonstrated poor short and long-term outcomes in patients with AMI and AF.^21^ The large Global Registry of Acute Coronary Events (GRACE) showed a three-fold increased risk of death for patients with new-onset AF during index hospitalization for acute coronary syndrome.^22^ This study concurs with previous findings and found AF as an independent predictor for increased odds of mortality while controlling for other known variables associated with increased mortality.

Several mechanisms are considered the possible etiology behind increased mortality in this sub-population. One theory suggested that due to shared risk factors between the two conditions, AF and CAD are both the products of these underlying risk factors that promote atrial ischemia, induce AF, and encourage obstructive coronary artery disease.^23^ Another hypothesis suggests reentry pathways with different refractory times create reentry circuits and, along with the structural remodeling of the left atrium, increase vulnerability to AF.^24, 25^ Very few cases of thromboembolic AMI have been reported with AF due to the associated systemic inflammatory state promoting prothrombotic environment.^26, 27^ Lastly, the increased ventricular rate can induce subendocardial ischemic and cause NSTEMI.^28^

One study found no association between a history of AF and significant CAD in patients undergoing coronary angiograms.^29^ In contrast, another research revealed that the vast majority of deaths in anticoagulated AF patients were from cardiac causes (37%) rather than stroke (9.8%).^30^ Adding complexity, a prospective cohort study demonstrated an association between AF and NSTEMI and not with STEMI.^31^ Despite AF showing an increased mortality in AMI,^20^ it remains uncertain whether increased coronary events are the primary cause.

Pooled data from a 2016 meta-analysis showed a 39% elevated risk of AMI in AF patients and a two-fold increased likelihood of cardiovascular events or death compared to patients without AF.^32^ In another longitudinal study, AF was associated with a two-fold increased risk for MI, and the risk of mortality was notably higher in an African American female on anticoagulation with a high ChadsVasc score.^13^ However, our study found no statistically significant difference in mortality between genders (adjusted OR 1.02, p 0.498).

Single-center retrospective analysis showed no association between anatomical characteristics of CAD and AF; however, it found more severe CAD in patients with AF.^33^ Given the high mortality linked with AF and AMI and previous studies indicating a higher likelihood of severe CAD in this patient population, data was analyzed in this study to assess the trend of ischemic evaluation and intervention in patients admitted with AMI and stratified them based on AF status. The study showed increased mortality and decreased odds of undergoing ischemic evaluation with a nuclear stress test or diagnostic coronary angiogram. The study also showed lesser odds of receiving percutaneous interventions.

Similarly, patients with AF were more likely to have serious cardiovascular complications such as VT, VF, cardiogenic shock, and cardiac arrest compared to patients without AF. The average length of hospital stays and total hospital charges were also high in AF. This study validates the previous findings of increased mortality in patients with CAD and concomitant AF. It further highlights the deficits in diagnostic evaluation in this patient population, which was not examined before. The study indicated an increased propensity for cardiac complications and elevated resource utilization, underscoring the crucial importance of treating AF as a distinct risk factor for CAD and advocating for early ischemic evaluation.

Limitations: Our study had the following limitations. It included all-cause mortality as the outcome instead of cardiovascular-specific mortality, as CV mortality is not available in the dataset. There is a potential for coding error as the ICD-10 codes’ accuracy depends on the treating physician. The patients with missing information were excluded from the analysis, which may infer selection bias. The anticoagulation status of AF patients was not known.

## Conclusion

This study examined the impact of AF on CAD mortality, cardiac complications, length of stay, and total hospital charges. AMI patients with AF had higher odds of mortality. They were less likely to undergo ischemic workup or intervention, more likely to suffer serious cardiac complications, and, on average, had longer lengths of hospital stay and higher hospital charges. AF appears to affect cardiovascular mortality independently. AF should be considered as a marker for underlying CAD. More research is required to understand the challenges of obtaining ischemic workup in AF patients. More education is needed to promote early consideration of ischemic evaluation in patients with AF.

## Data Availability

HCUP de-identified protected data.

## Non-standard abbreviations and acronyms

AMI: Acute myocardial infarction

AF: Atrial fibrillation

STEMI: ST-elevation myocardial infarction

NSTEMI: Non-ST elevation myocardial infarction

VT: Ventricular tachycardia

VF: Ventricular fibrillation

LOS: length of stay

